# Pre-morbid risk factors for amyotrophic lateral sclerosis: prospective cohort study

**DOI:** 10.1101/2021.07.01.21259861

**Authors:** G. David Batty, Catharine R. Gale

**Author notes:** Correspondence:* David Batty, Department of Epidemiology and Public Health, University College London, 1-19 Torrington Place, London, UK, WC1E 6BT. E. Access to data:* Data from UK Biobank (http://www.ukbiobank.ac.uk/) are available to *bona fide* researchers upon application. This research has been conducted under application 10279.

## Abstract

**Background:** As a neurodegenerative disorder with high case-fatality, there is a need to identify primary, modifiable risk factors for amyotrophic lateral sclerosis (ALS). Their detection has, however, proved elusive and this may be ascribed to the scarcity of well-characterised, sufficiently-powered cohort studies necessary to explore the aetiology of this rare condition.

**Methods:** UK Biobank is an on-going, closed, prospective cohort study in which 502,524 participants (273,420 women) have been linked to national hospital and mortality registries. Baseline data collection took place between 2006 and 2010 when a range of psychosocial, physiological, and demographic data were captured.

**Results:** Approximately 11 years of event surveillance gave rise to 301 hospitalisations and 279 deaths ascribed to ALS. After left censoring to account for reverse causality and adjustment for confounding factors, being older (hazard ratio per 10 year increase; 95% confidence interval: 1.92; 1.58, 2.33) and male (1.37; 1.00, 1.87) were associated with elevated rates of hospitalisation for ALS. Similar effects were apparent when death due to the disorder was the outcome of interest. Of the remaining 23 potential risk indices, however, there was only a suggestion that taller people experienced an increased risk of hospitalisation (per SD increase: 1.31; 1.09, 1.59).

**Conclusions:** In the present study, a comprehensive array of potential risk indices were essentially unrelated to later ALS.

## Introduction

Amyotrophic lateral sclerosis (ALS), also known as motor neuron disease, involves the unabated degeneration of nerve cells responsible for voluntary muscle movement. With there being no effective treatment, in most cases, death, typically from respiratory failure, commonly occurs within 3 years of symptom emergence.^1,2^ This brings into sharp focus the need for primary prevention research.

While marked geographical differentials in case occurrence^3^ broadly implicates both genetic and non-genetic causes of ALS, a secular increase in rates over three decades years^4^ – too rapid to be explained by changes in the gene pool – provides some circumstantial support for a role of environmental determinants. There is strong evidence to suggest being male and older are associated with an elevation in risk,^5^ however, studies examining the role of modifiable environmental characteristics report inconsistent and often counter-intuitive results. Thus, although higher pre-diagnostic body weight appears to confer protection against ALS,^6,7^ cigarette smoking has revealed discordant associations,^8,9^ as have physical activity,^10^ alcohol intake,^11,12^ socioeconomic status,^13,14^ and physical comorbidities such as diabetes,^15,16^ amongst many other factors.^17^

The heterogeneity of these findings may, at least in part, be explained by the methodological shortcomings of selected studies. Given the rarity of ALS, the most time- and cost-efficient method for identifying risk factors is the case-control study, interpretation of findings from which is complicated by recall bias and, potentially, reverse causality, such that biomedical indices are captured in the presence of the disorder. Accordingly, we used a well-characterised cohort study of 500,000 people with on-going surveillance for mortality and morbidity from ALS in order to better understand the role, if any, of a range of modifiable biological, psychosocial, and behavioural characteristics. In particular, our purpose was to further examine some extant associations and also to relate previously untested characteristics to the onset of the disorder. There are, for example, reasons to anticipate associations between ALS and earlier measurements of cognitive function, psychological distress, and lung function, not least because these characteristics may be pre-clinical features of the disorder.^18^ These indices have also shown prognostic value in studies of patient groups – whereby less favourable levels are associated with adverse outcomes including disability severity and reduced life expectancy^19,20^ – they are untested as risk factors for disease onset.

## Methods

UK Biobank, a UK-wide closed, prospective cohort study, has been described in detail.^5^ In brief, between 2006 and 2010, 502,649 participants aged 37 to 73 years attended 22 geographically disparate research clinics when they completed a questionnaire, underwent an interview, and took part in various physical assessments. Ethical approval was obtained from the National Health Service National Research Ethics Service with all participants providing written informed consent. Using fully anonymised data, the present analyses did not require additional permissions. We followed STROBE guidelines for the presentation of original epidemiological research.^21^

### Assessment of baseline characteristics

Ethnicity was self-reported and categorised as White, Asian, Black, Chinese, Mixed, or other ethnic group.^22^ A social isolation scale was derived from enquiries concerning number of people in household, visits with friends/family, and social activities.^23^ Cigarette smoking, physical activity, and alcohol consumption were measured using standard enquiries.^24^ Height and weight were recorded directly and body mass index was calculated using the usual formula (weight, kg/height,^2^ m^2^); overweight and obesity were denoted by values ≥25kg/m^2^.^25^ Self-reported physician diagnosis was collected for ALS, vascular or heart problems, diabetes, and cancer. We defined hypertension according to existing guidelines: measured systolic/diastolic blood pressure ≥140/90 mmHg (two assessments) and/or self-reported use of antihypertensive medication.^26,27^

Forced expiratory volume in one second, a measure of pulmonary function, was quantified using spirometry with the best of three technically satisfactory exhalations used in our analyses. Handgrip strength was measured using a hydraulic hand dynamometer with the participant maximally squeezing the handle of the dynamometer while seated for 3 seconds; an average of readings from the right and left hand was used. Non-fasting venous blood, available in a sub-sample, was drawn with assaying conducted at a dedicated central laboratory for high-density lipoprotein cholesterol and glycated haemoglobin A1c.^28^

Study members were asked if they had ever been under the care of a psychiatrist for any mental health problem.^29^ Symptoms of psychological distress – anxiety, worrying, anhedonia, and depression – were measured using the four item version of the Patient Health Questionnaire (PHQ-4)^30,31^ in which individual items are rated on a Likert scale from 0 (“not at all”) to 3 (“nearly every day”) such that total scores range from 0 to 12 (higher scores denote greater distress). A score of 3 or above was used to indicate high distress in the present analyses.^29^ Neuroticism was measured with the 12-item Eysenck Personality Questionnaire-Revised Short Form;^32,33^ again, higher scores denote higher levels. Verbal and numerical reasoning was captured using a computerized 13-item multiple-choice test with a two-minute time limit. The score was the number of correct answers.^34^ This test was introduced after the beginning of the baseline assessment period so data are available for a subset of study members (N=180,914).^35,36^ Reaction time was measured using a computerized Go/No-Go “Snap” game.^36^ Participants were presented with electronic images of two cards. If symbols on the cards were identical, participants were instructed to immediately push the button-box using their dominant hand. The first five pairs were used as a practice with the remaining seven pairs, containing four identical cards, forming the assessment. Reaction time score was the mean time to press the button when each of these four pairs was presented. Choice reaction time correlates strongly with single mental tests that involve complex reasoning and knowledge.^37^

For educational qualifications, we used a two category variable (college/university degree versus lower). Total annual household income before tax was self-reported and classified into three groups (<18,000 versus higher). The Townsend deprivation index was our indicator of neighbourhood socioeconomic circumstances; a continuously scored variable, higher values denote greater deprivation.^35^

### Ascertainment of amyotrophic lateral sclerosis cases

Study participants were linked to the National Health Service’s Central Registry which provided vital status data on study members and, where applicable, cause of death. Linkage was also made to hospital in-patients records via the Hospital Episode Statistics, a registry of all hospitalisations in the UK.^38^ Using both databases, ALS was denoted by International Classification of Disease code G122.

### Derivation of analytical sample and statistical analyses

To addresses concerns regarding reverse causality – the notion that ALS might influence the exposure of interest (e.g., physical activity, lung function) rather than the converse – we excluded 56 people who self-reported ALS at baseline medical examination. Additionally, to capture study members with potentially subclinical (undiagnosed) ALS, we left-censored study members such that those who were hospitalised for or died (N=72) from the condition within the first 3 years of baseline were also excluded. This resulted in an analytical sample of 502,599 people (273,454 women). To summarise risk factor–ALS associations, we computed hazard ratio with accompanying 95% confidence intervals.^39^ In these analyses, we used calendar period as the time scale and study members were censored at date of hospitalisation or death from ALS, or end of follow-up (4^th^ October 2020 for mortality, 24^th^ November 2020 for hospitalisation) – whichever came first.

## Results

Event surveillance gave rise to 301 hospitalisations (mean 11.8 years of follow-up) and 279 deaths (11.4 years) due to ALS. In tables 1 (hospitalisations) and 2 (deaths) we show the relationships between twenty-five baseline characteristics and ALS outcomes. After adjustment for selected confounding factors, being older (hazard ratio per 10 year increase; 95% confidence interval: 1.92; 1.58, 2.33) and male (1.37; 1.00, 1.87) were associated with elevated rates of hospitalisation for ALS. Similar effects were apparent when death ascribed to the disorder was the outcome of interest. Of the remaining 23 potential risk indices, however, there was only a suggestion that taller people experienced an increased risk of hospitalisation (per SD increase: 1.31; 1.09, 1.59); weaker effects were apparent in the mortality analyses (1.21; 0.98, 1.48). While there was some indication of an elevated rate of hospitalisations in people who reported cigarette smoking, confidence intervals included unity and there was marked attenuation after adjustment for covariates which included socioeconomic status and health behaviours (1.10; 0.73, 1.66).

**Table 1.** Hazard ratios for the association between baseline characteristics and hospitalisation for amyotrophic lateral sclerosis, UK Biobank 2006 to 2020.

|  | Age- and sex-adjusted |  | Multiply-adjusted |  |
| --- | --- | --- | --- | --- |
|  | N events /<br>N at risk | Hazard ratio<br>(95% confidence<br>interval) | N events /<br>N at risk | Hazard ratio<br>(95% confidence<br>interval) |
| **Demographic factors** |  |  |  |  |
| Age (yrs), per 10 year increase | 301/502,524 | 1.96 (1.67, 2.31) | 258/446,321 | 1.92 (1.58, 2.33) |
| Male | 301/502,524 | 1.23 (0.98, 1.54) | 258/446,321 | 1.37 (1.00, 1.87) |
| Non-white ethnicity | 300/499,634 | 0.65 (0.32, 1.31) | 258/446,321 | 0.42 (0.17, 1.04) |
| Socially isolated | 301/502,524 | 0.91 (0.60, 1.36) | 258/446,321 | 0.82 (0.52, 1.31) |
| **Lifestyle factors** |  |  |  |  |
| Current smoker | 300/499,571 | 1.71 (0.81, 1.68) | 258/446,321 | 1.10 (0.73, 1.66) |
| No physical activity | 298/495,378 | 0.89 (0.55, 1.45) | 258/446,321 | 0.88 (0.51, 1.53) |
| Drinks alcohol daily/almost daily | 300/501,021 | 0.87 (0.65, 1.15) | 257/446,013 | 0.89 (0.66, 1.22) |
| Obese/overweight | 296/499,469 | 0.80 (0.62, 1.03) | 257/446,013 | 0.86 (0.65, 1.14) |
| **Comorbidities** |  |  |  |  |
| Vascular or heart disease | 300/500,300 | 1.13 (0.89, 1.44) | 257/445,599 | 1.08 (0.83, 1.42) |
| Hypertension | 296/493,912 | 0.99 (0.77, 1.27) | 256/443,514 | 1.03 (0.78, 1.35) |
| Diabetes | 299/499,907 | 1.10 (0.70, 1.72) | 258/446,321 | 1.10 (0.67, 1.82) |
| Cancer | 200/499,749 | 1.17 (0.81, 1.68) | 257/445,179 | 1.27 (0.85, 1.88) |
| **Biomarkers** |  |  |  |  |
| Lung function, per SD (0.8 L) increase | 298/453,787 | 0.91 (0.74, 1.11) | 258/446,321 | 0.92 (0.77, 1.09) |
| Hand grip strength, per SD (11.3 kg) increase | 299/500,164 | 0.97 (0.82, 1.16) | 258/445,857 | 1.03 (0.84, 1.25) |
| High-density lipoprotein, 25<sup>th</sup> vs 75<sup>th</sup> centile | 262/429,780 | 1.16 (0.81, 1.68) | 224/385,554 | 1.18 (0.78, 1.77) |
| HbA1C, 25<sup>th</sup> vs 75<sup>th</sup> centile | 279/466,404 | 0.92 (0.65, 1.30) | 243/418,663 | 1.10 (0.73, 1.64) |
| Height, per SD (9.28 cm) increase | 296/499,990 | 1.17 (0.99, 1.38) | 257/445,898 | 1.31 (1.09, 1.59) |
| **Psychological factors** |  |  |  |  |
| Psychiatric consultation | 300/498,839 | 1.12 (0.79, 1.58) | 257/444,419 | 1.03 (0.70, 1.52) |
| High psychological distress | 267/448,959 | 1.25 (0.94, 1.65) | 232/401,583 | 1.27 (0.93, 1.72) |
| Neuroticism, per SD (3.28 points) increase | 154/425,652 | 1.03 (0.88, 1.21) | 132/381,324 | 1.03 (0.86, 1.23) |
| Reasoning, per SD (2.16 points) increase | 120/180,869 | 0.78 (0.64, 0.93) | 103/162,497 | 0.85 (0.69, 1.05) |
| Reaction time, per SD (118.2 ms) increase | 299/496,751 | 1.09 (0.98, 1.21) | 256/442,583 | 1.06 (0.94, 1.19) |
| **Socioeconomic factors** |  |  |  |  |
| No college/university degree | 293/492,384 | 1.11 (0.86, 1.44) | 254/442,098 | 1.15 (0.87, 1.54) |
| Low annual household income | 250/425,341 | 0.99 (0.74, 1.31) | 218/383,913 | 0.88 (0.64, 1.23) |
| Deprivation score, per point disadvantage | 299/501,897 | 1.01 (0.97, 1.05) | 258/446,321 | 1.01 (0.97, 1.06) |

Multiple adjustment is adjustment for age, sex, ethnicity, smoking status, physical activity, diabetes, lung function, and deprivation score

**Table 2.** Hazard ratios for the association between baseline characteristics and death from amyotrophic lateral sclerosis, UK Biobank 2006 to 2020.

|  | Age- and sex-adjusted |  | Multiply-adjusted |  |
| --- | --- | --- | --- | --- |
|  | N events /<br>N at risk | Hazard ratio (95%<br>confidence<br>interval) | N events /<br>N at risk | Hazard ratio (95%<br>confidence<br>interval) |
| **Demographic factors** |  |  |  |  |
| Age (yrs), per 10 year increase | 279/502,524 | 2.42 (2.02, 2.90) | 236/446,321 | 2.41 (1.95, 2.99) |
| Male | 279/502,524 | 1.28 (1.01, 1.62) | 236/446,321 | 1.25 (0.90, 1.12) |
| Non-white ethnicity | 278/499,634 | 0.46 (0.19, 1.12) | 236/446,321 | 0.33 (0.10, 1.04) |
| Socially isolated | 279/502,524 | 0.94 (0.61, 1.43) | 236/446,321 | 0.84 (0.51, 1.37) |
| **Lifestyle factors** |  |  |  |  |
| Current smoker | 277/499,571 | 1.16 (0.79, 1.72) | 236/446,321 | 1.14 (0.73, 1.77) |
| No physical activity | 274/495,378 | 0.98 (0.59, 1.62) | 236/446,321 | 1.09 (0.63, 1.89) |
| Drinks alcohol daily/almost daily | 278/501,021 | 1.10 (0.84, 1.46) | 235/446,013 | 1.20 (0.89, 1.61) |
| Obese/overweight | 277/499,469 | 0.90 (0.69, 1.18) | 235/445,540 | 0.92 (0.68, 1.23) |
| **Comorbidities** |  |  |  |  |
| Vascular or heart disease | 278/500,300 | 1.09 (0.85, 1.40) | 235/445,599 | 1.12 (0.85, 1.48) |
| Hypertension | 274/493,912 | 1.02 (0.78, 1.32) | 235/443,514 | 1.11 (0.84, 1.49) |
| Diabetes | 278/499,907 | 0.91 (0.55, 1.50) | 236/445,321 | 1.02 (0.59, 1.76) |
| Cancer | 277/499,749 | 1.02 (0.68, 1.54) | 234/445,179 | 0.93 (0.58, 1.49) |
| **Biomarkers** |  |  |  |  |
| Lung function, per SD (0.8 L) increase | 242/453,729 | 1.03 (0.87, 1.22) | 236/446,321 | 1.03 (0.86, 1.23) |
| Hand grip strength, per SD (11.3 kg) increase | 278/500,164 | 1.02 (0.85, 1.22) | 236/445,857 | 0.99 (0.81, 1.22) |
| High-density lipoprotein, 25<sup>th</sup> vs 75<sup>th</sup> centile | 237/429,780 | 1.06 (0.72, 1.56) | 200/385,554 | 1.02 (0.66, 1.57) |
| HbA1C, 25<sup>th</sup> vs 75<sup>th</sup> centile | 255/466,404 | 0.90 (0.62, 1.32) | 220/418,663 | 1.05 (0.68, 1.61) |
| Height, per SD (9.28 cm) increase | 277/499,990 | 1.17 (0.99, 1.39) | 235/445,898 | 1.21 (0.98, 1.48) |
| **Psychological factors** |  |  |  |  |
| Psychiatric consultation | 277/498,839 | 0.85 (0.57, 1.27) | 235/444,419 | 0.81 (0.52, 1.27) |
| High psychological distress | 244/448,959 | 0.99 (0.72, 1.36) | 210/401,583 | 0.98 (0.69, 1.39) |
| Neuroticism, per SD (3.28 points) increase | 100/425,652 | 0.93 (0.76, 1.15) | 83/381,324 | 0.86 (0.68, 1.09) |
| Reasoning, per SD (2.16 points) increase | 88/180,869 | 0.82 (0.66, 1.02) | 74/162,497 | 0.86 (0.67, 1.09) |
| Reaction time, per SD (118.2 ms) increase | 275/496,751 | 0.97 (0.86, 1.10) | 232/442,583 | 0.98 (0.86, 1.13) |
| **Socioeconomic factors** |  |  |  |  |
| No college/university degree | 272/492,384 | 1.09 (0.83, 1.42) | 233/442,098 | 1.01 (0.75, 1.35) |
| Low annual household income | 227/425,341 | 0.96 (0.71, 1.29) | 199/383,913 | 0.88 (0.62, 1.24) |
| Deprivation score, per point disadvantage | 278/501,897 | 0.99 (0.95, 1.03) | 236/446,321 | 1.00 (0.96, 1.05) |

Multiple adjustment is adjustment for age, sex, ethnicity, smoking status, physical activity, diabetes, lung function, and deprivation score

## Discussion

The main finding of the present study was that, aside of older age, being male, and greater physical stature, there was no clear evidence of an association between ALS and the twenty-five risk indicators. The exceptions were the two most established risk factors – age and sex; that these associations were recapitulated here gives us some confidence in the generally negative results for the remainder of the baseline characteristics. These null findings are in keeping with observations from systematic reviews of risk factors for ALS^17,20,40,41^ and those for other neurodegenerative disorders such as dementia.^42-44^

The positive association between height and ALS – also apparent for cancer^45,46^ – potentially implicates early life exposures in the occurrence of this condition. The same has been speculated for dementia,^47^ although a reverse height gradient to that seen herein has been reported.^48^ While under a degree of genetic control, relative to their shorter counterparts, taller people have, on average, have been exposed to a more favourable pre-adult environment, which includes an optimal diet, fewer bouts of illness, more affluent social circumstances, and a low stress household,^49^ though we are unable to identify which, if any, of these correlates may be exerting an influence on ALS risk.

### Study strengths and weaknesses

The present study has its strengths, including its size which facilitates the accumulation of a sufficiently high number of cases for analyses of a rare health outcome alongside left-censoring to take into account reverse causality, and the well characterised nature of the study participants. Further, in the aetiological literature, investigators often focus on the predictive value of a single characteristic rather than take a comprehensive approach, as we have, so facilitating cross-comparison. Inevitably, however, our work has several weaknesses. First, the present study sample comprises only the 5.5% of the target population who agreed to participate.^50^ As has been demonstrated,^51,52^ the data material is therefore inappropriate for estimation of risk factor or disease prevalence and for event incidence, including ALS. These observations do not, however, seem to influence reproducibility of the association of established risk factors for non-communicable disease such as vascular disease and selected cancers, and other health endpoints such as suicide and selected cancers.^52^ We think the same reasoning can be applied to the present analyses for ALS.

Second, while we set out to further test the relation of selected risk factors (e.g., cigarette smoking, diabetes), and potentially identify novel ones (e.g., cognitive function, psychological distress), several other factors (e.g., alcohol intake, extant cancer, blood lipids) were not hypothesis-driven. Lastly, baseline data are certainly time-varying in the period between study induction and end of follow-up. This is a perennial issue in cohort studies and one we were able to investigate using data from a resurvey that took place around 8 years after baseline examination in a representative sub-sample of participants.^36,53^ Analyses revealed moderate to high stability for some covariates, including education (r=0.86, p<0.001, N=30,350) and body mass index (r=0.90, p<0.001, N=34,662), whereas the magnitude was somewhat lower for co-morbidities such as diabetes (r=0.63, P<0.001, N= 31,037) and serious mental illness (r=0.64, p<0.001, N=47,291), as it was for cigarette smoking (r=0.60, p<0.001, N=31,037).

In conclusion, other than the known associations for age and sex, we did not find convincing evidence of links with ALS for other risk indices in the present analyses. A role for height perhaps warrants further enquiry.

## Data Availability

Data from UK Biobank (http://www.ukbiobank.ac.uk/) are available to bona fide researchers upon application.

## Acknowledgements

We thank the participants in UK Biobank for their forbearance.

